# The behavioral variant of Alzheimer’s disease does not show a selective loss of Von Economo and phylogenetically related neurons in the anterior cingulate cortex

**DOI:** 10.1101/2021.10.30.21265649

**Authors:** E.H. Singleton, Y.A.L Pijnenburg, P. Gami-Patel, B.D.C. Boon, F. Bouwman, J. Papma, H. Seelaar, P. Scheltens, L.T. Grinberg, S. Spina, A.L. Nana, G.D. Rabinovici, W.W. Seeley, R. Ossenkoppele, A.A. Dijkstra

**Affiliations:** Alzheimer Center Amsterdam, Department of Neurology, Amsterdam Neuroscience, Vrije Universiteit Amsterdam, Amsterdam UMC, Amsterdam, the Netherlands; Department of Pathology, Amsterdam Neuroscience, Vrije Universiteit Amsterdam, Amsterdam UMC, Amsterdam, the Netherlands; Neurology, Erasmus University Medical Center, Rotterdam, the Netherlands; Netherlands; Radiology, Erasmus University Medical Center, Rotterdam, the Netherlands; Netherlands; Departments of Pathology, University of California San Fransisco, San Francisco, USA; Departments of Neurology, University of California San Francisco, San Francisco, USA; Radiology and Biomedical Imaging, University of California San Francisco, San Francisco, USA; Clinical Memory Research Unit, Department of Clinical Sciences Malmö, Lund University, Sweden

## Abstract

**Background:** The neurobiological origins of the early and predominant behavioral changes seen in the behavioral variant of Alzheimer’s disease (bvAD) remain unclear. A selective loss of Von Economo Neurons (VENs) and phylogenetically related neurons have been observed in behavioral variant frontotemporal dementia (bvFTD) and several psychiatric diseases. Here, we assessed whether these specific neuronal populations show a selective loss in bvAD.

**Methods:** VENs and GABA receptor subunit theta (GABRQ)-immunoreactive pyramidal neurons of the anterior cingulate cortex (ACC) were quantified in post-mortem tissue of patients with bvAD (n=9) and compared to typical AD (tAD, n=6), bvFTD due to frontotemporal lobar degeneration based on TDP-43 pathology (FTLD, n=18) and controls (n=13) using ANCOVAs adjusted for age and Bonferroni correceted. In addition, ratios of VENs and GABRQ-immunoreactive (GABRQ-ir) pyramidal neurons over all Layer 5 neurons were compared between groups to correct for overall Layer 5 neuronal loss.

**Results:** The number of VENs or GABRQ-ir neurons did not differ significantly between bvAD (VENs: 26.0±15.3, GABRQ-ir pyramidal: 260.44±87.13) and tAD (VENs: 32.0±18.1, *p*=1.00, GABRQ-ir pyramidal: 349.83±109.64, *p*=0.38) and controls (VENs: 33.5±20.3, *p*=1.00, GABRQ-ir pyramidal: 339.38±95.88, *p*=0.37). Compared to bvFTD, patients with bvAD showed significantly more GABRQ-ir pyramidal neurons (bvFTD: 140.39±82.58, *p*=0.01) and no significant differences in number of VENs (bvFTD: 10.9±13.8, *p*=0.13). Results were similar when assessing the number of VENs and GABRQ-ir relative to all neurons of Layer 5.

**Discussion:** VENs and phylogenetically related neurons did not show a selective loss in the ACC in patients with bvAD. Our results suggest that, unlike in bvFTD, the clinical presentation in bvAD may not be related to the loss of VENs and related neurons in the ACC.

## Introduction

Von Economo Neurons (VENs) constitute a specialized type of large bipolar projection neurons, located in Layer 5 of the anterior cingulate cortex (ACC) and frontoinsular cortex (FI) [1] and have been associated with behavioral dysregulation in bvFTD [2-4], autism [5, 6] and schizophrenia [7, 8]. These neurons are part of a larger neuronal population in the same cortical layer, the GABA_A_ receptor subunit theta (GABRQ) immunoreactive neurons [9]. In addition to VENs, the GABRQ neuronal population in the ACC is also reduced in bvFTD [10] and schizophrenia [11]. The behavioral variant of Alzheimer’s disease (bvAD) shows clinical overlap with behavioral variant of frontotemporal dementia (bvFTD), characterized by early and predominantly behavioral changes, such as disinhibition, apathy and compulsiveness, with AD as underlying pathology [12-15]. Previous investigations showed conflicting results regarding predominant involvement of anterior regions that are essential for socio-emotional functioning in terms of atrophy [12, 16-19], hypometabolism [20-23] or and tau pathology [19, 24-27]. In the absence of a clear neuroanatomical origin, an alternative explanation may not lie in topographical spreading of AD pathological processes, but instead in a selective loss of specialized neurons regulating social behavior. A previous study showed no selective loss of VENs in the ACC in bvAD with substantial Lewy body pathology[28], leaving the role of VENs in “pure” bvAD unknown. Furthermore, the phylogenetically realted neurons have not been assessed in bvAD. Here, we studied the number of VENs and GABRQ-immunoreactive (GABRQ-ir) neurons in carefully phenotyped bvAD cases compared to patients with typical AD (tAD), bvFTD and controls. We hypothesized that bvAD would show comparable reductions in numbers and ratios of VENs and phylogenetically related neurons with bvFTD, and greater reductions compared to controls and tAD patients.

## Methods

### Participants

Post-mortem brain tissue was obtained from the University of California San Francisco Neurodegenerative Disease Brain Bank (UCSF NDBB) for 3 bvAD cases and the remaining cases were obtained from the Netherlands Brain Bank (NBB; Amsterdam, The Netherlands, https://www.brainbank.nl). Participants and their caregivers provided informed consent to undergo autopsy according to the Declaration of Helsinki, and all study procedures were approved by the institutional review boards at the participating sites. Extensive clinical reports were available from all donors. Patients with bvAD were included if they showed a) a primary pathological diagnosis of AD, b) met during life ≥2 of 6 core clinical criteria for bvFTD [29], consisting of early and predominant apathy, loss of empathy, disinhibition, compulsive behaviors, hyperorality and dysexecutive functioning based on patient chart reviews (reviewed by AAD, EHS, RO; see Table S1). We included six donors with a typical amnestic presentation of AD (tAD) [30] and primary pathological diagnosis of AD [31] and 18 donors with bvFTD [29] with underlying FTLD-TDP pathology, consisting of 9 cases with a *C9orf72* repeat expansion, 5 cases with progranulin mutations and 4 cases with a sporadic origin (n=1 TDP-E, n=1 TDP-C, n=2 TDP-A), according to FTLD consensus criteria [32] as a reference group. In addition, we included 13 cognitively unimpaired individuals (Table 1 and Table S1). Donors were included prospectively, with as much age matching as possible, with inherent differences in age between typical AD and bvAD and bvFTD cases. Donors with significant concomitant pathology leading to mixed pathological diagnoses were excluded from the study; both bvAD and tAD donors were restricted to early-stage PD Lewy body scores (Braak 0-3, or amygdala only) [33] and limbic-only TDP-43 aggregation [34] (Table S3).

**Table 1.**
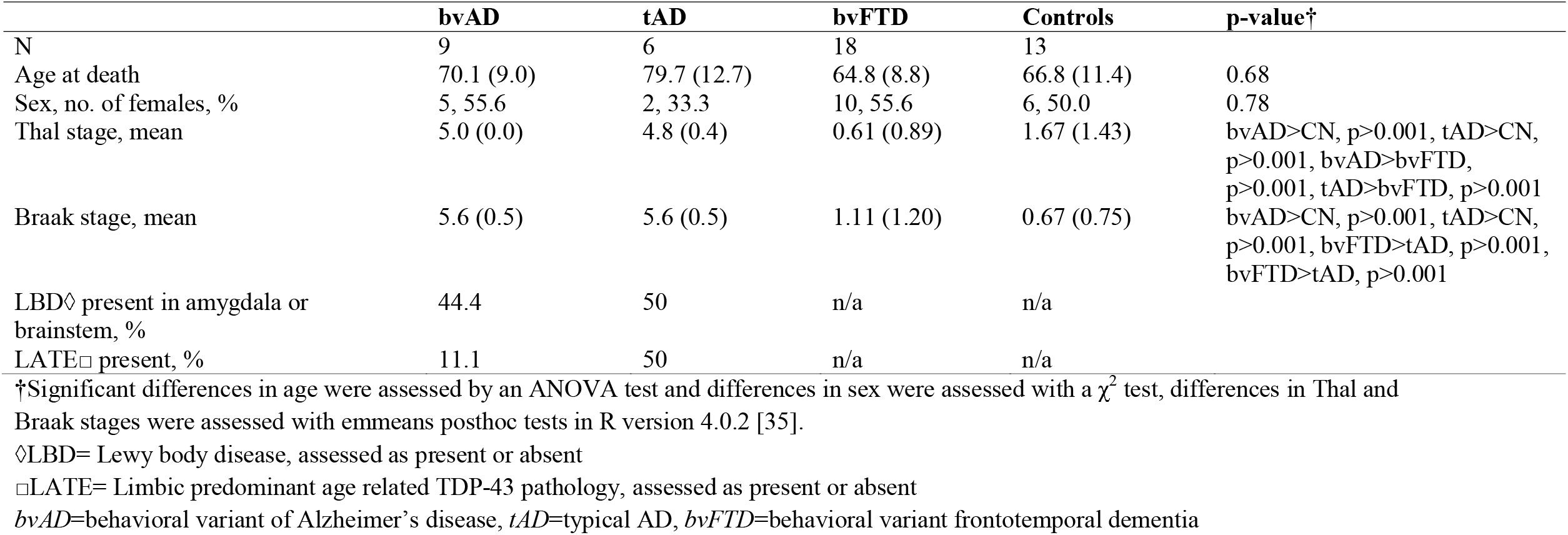
Demographic and pathological characteristics of participants by diagnostic group

### Immunohistochemical procedures

Immunohistochemical procedures were performed as previously reported [10]. Briefly, sequential 10 μm sections of the ACC adjacent to the genu were sampled from the right hemisphere and stained for GABA receptor subunit epsilon (GABRE; 1:1000; HPA045918, Sigma Aldrich, St. Louis, MO) and GABRQ (1:750; HPA002063; Sigma Aldrich) and counterstained with haematoxylin as previously reported [10]. Quantification was performed using the Meander option in Stereoinvestigator, and GABRE staining was used to outline Layer 5a and GABRQ staining to visualize VENs and related neurons. After delineation of Layer 5, two blinded raters (PGP & AAD) classified each pyramidal neuron and VEN in Layer 5 based on morphology into four groups: GABRQ-ir VENs, GABRQ-ir pyramidal neurons, GABRQ-negative VENs and GABRQ-negative neurons. Previous work performed by the same raters in a similar sample showed interrater reliability of α=0.06 (REF). VENs were distinguished from pyramidal neurons by their large bipolar cell body and thick dendrites [1]. Finally, ratios relative to all Layer 5 neurons were calculated to control for varying size of the ACC and overall neurodegeneration. A step-by-step illustration of these procedures is provided in Figure 1.

**Figure 1.**
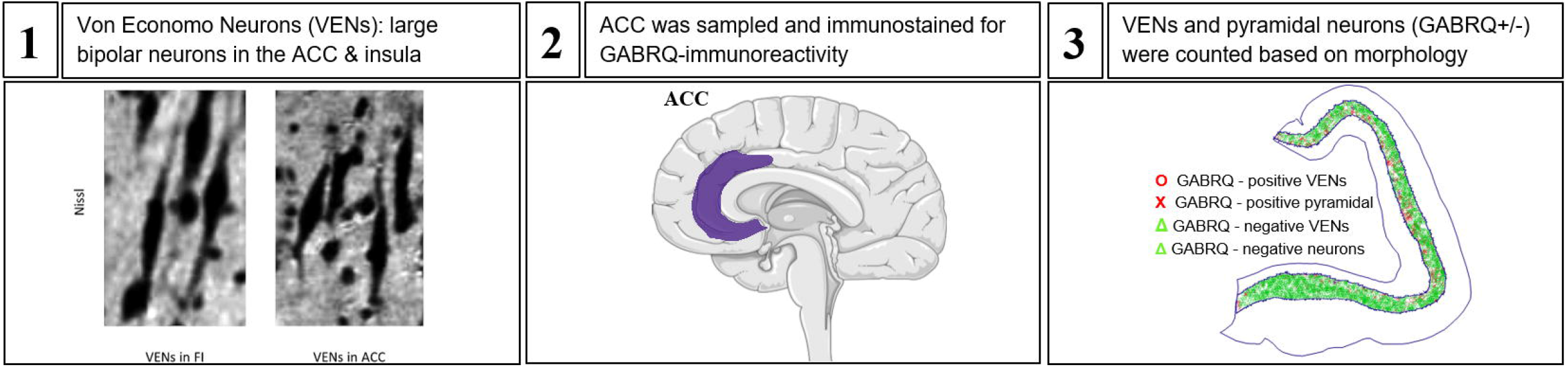
Schematic overview of methodology. **Panel 1** shows an illustration of VENs, adapted from Banovac et al. 2021 [56]. **Panel 2** shows the location of the ACC in the human cortex. Both images were obtained from smartservier.com. **Panel 3** shows an illustration of the type of neurons in the ACC after staining and counting.

### Statistical analyses

Differences in demographic characteristics were studied using ANOVAs or χ^2^ tests where appropriate. Main effects in the number of VENs, GABRQ-ir pyramidal neurons and the ratios VENs/all neurons and GABRQ-ir pyramidal/all neurons were analyzed using ANCOVAs adjusting for age and between group differences were assessed using emmeans post-hoc tests, adjusting for age, using a Bonferroni correction in R version 4.0.2 [35].

## Results

### Participant characteristics

Patient characteristics are provided in Table 1, Table S1 and Table S2. Mean age at death was 70.1±9.0 in cases with bvAD compared to 79.7±12.7 in tAD, 64.8±8.8 in bvFTD and 66.8±11.4 in controls, the proportion of females was 55.6% in bvAD compared to 33.3% in tAD, 55.6% in bvFTD and 50.0% in controls.

### Copathologies across groups

There were no significant differences in age and sex among groups (*p*=0.68 and *p*=0.78, respectively). Lewy body copathology in the brainstem and/or amygdala was present in 44% of bvAD and 40% tAD cases and TDP copathology in the limbic regions was present in 11% bvAD and 40% of tAD cases. In addition, cerebral amyloid angiopathy was present in 89% of bvAD and 80% of tAD cases, cerebrovascular disease in 63% of bvAD and 80% of typical AD cases and aging-related tau astrogliopathy (ARTAG) was present in 56% of bvAD and 80% of typical AD cases.

### VENs and GABRQ-immunoreactive neurons across groups

The numbers and ratios of VENs (GABRQ-ir and –negative) and GABRQ-ir pyramidal neurons are shown in Figure 2 and Table S4. After adjustment for age, there was a significant difference between the groups in the number of VENs, *F*(1,43)=12.6, *p*<0.001, the number of GABRQ-ir pyramidal neurons, *F*(1,43)=25.2, *p*<0.0001, the ratio VENs/all neurons, *F*(1,43)=9.0, *p*=0.004, the ratio GABRQ-ir pyramidal/all neurons, *F*(1,43)=14.9, *p*<0.001. Post-hoc tests, adjusted for age, showed no significant differences in the number of VENs or GABRQ-ir pyramidal neurons in bvAD (VENs: 26.0±15.3, GABRQ-ir pyramidal: 260.44±87.13) compared to tAD (VENs: 32.0±18.1, *p*=1.00, GABRQ-ir pyramidal: 349.83±109.64, *p*=0.38) and controls (VENs: 33.5±20.3, *p*=1.00, GABRQ-ir pyramidal: 339.38±95.88, *p*=0.37). In order to correct for varying size of the ACC, ratios relative to all Layer 5 neurons of the ACC were analyzed. The ratios of VENs and GABRQ-ir pyramidal vs all neurons showed a similar pattern, with no significant differences between bvAD (ratio VENs/all neurons: 0.009±0.004, ratio GABRQ-ir pyramidal/all neurons: 0.098±0.019) compared to tAD (ratio VENs/all neurons: 0.012±0.005, *p*=1.00, ratio GABRQ-ir pyramidal/all neurons: 0.133±0.036, *p*=0.37) and compared to controls (ratio VENs/all neurons: 0.010±0.005, *p*=1.00, ratio GABRQ-ir pyramidal/all neurons: 0.106±0.029, *p*=1.00). As reported before [10], in the current study FTLD-TDP donors showed lower numbers of VENs (bvFTD: 10.9±13.8), GABRQ-ir pyramidal neurons (bvFTD: 140.39±82.58) and ratios VENs/all neurons (bvFTD: 0.005±0.005) and GABRQ-ir pyramidal/all neurons (bvFTD: 0.059±0.028) compared to controls (VENs, *p*=0.003, GABRQ-ir pyramidal, *p*<0.0001, ratio VENs/all neurons, *p*=0.02, ratio GABRQ-ir pyramidal/all neurons, *p*<0.001) and tAD (VENs, *p=*0.03, GABRQ-ir pyramidal, *p*<0.001, ratio VENs/all neurons: *p*=0.07, ratio GABRQ-ir pyramidal/all neurons: *p*<0.001). Here, we show lower though non-significantly different number of VENs (bvFTD: 10.9±13.8, *p*=0.13), GABRQ-ir pyramidal neurons (bvFTD: 150.2±91.5, *p*=0.009), and ratios of VENs/all neurons (bvFTD: 0.005±0.005, *p*=0.23) and significantly lower ratios of GABRQ-ir pyramidal/all neurons (bvFTD: 0.063±0.031, *p*=0.01) in FTLD-TDP donors than bvAD. Assessment of all subcategories, i.e. the number of GABRQ-ir VENs, GABRQ-negative VENs, GABRQ-ir pyramidal and GABRQ-negative pyramidal are shown in Table S2 and Figure S1. There was no association between the number of bvFTD symptoms and number of VENs or GABRQ-ir pyramidal neurons in bvAD cases (p=0.92 and p=0.97 resp, see Figure S3).

**Figure 2.**
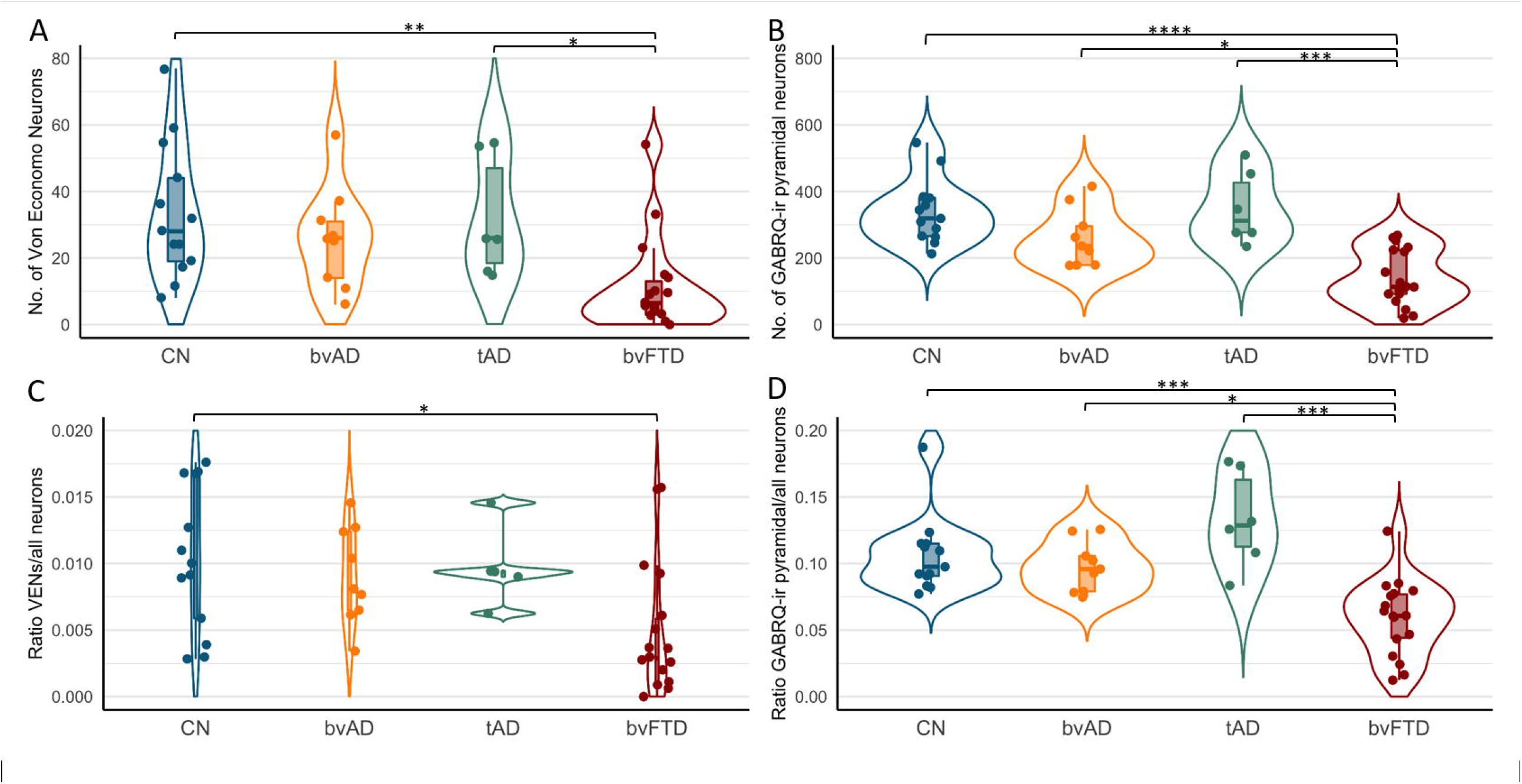
Numbers and ratios of VENs and GABRQ-immunoreactive pyramidal neurons in bvAD compared to tAD, bvFTD and controls **Panel A** shows the number of VENs counted based on GABRQ-immunohistochemistry across groups, representing the total number of GABRQ-immunopositive VENs and GABRQ-immunonegative VENs, showing no significant differences between bvAD, typical AD and controls. **Panel B** shows the number of GABRQ-immunoreactive pyramidal neurons) across groups, showing no significant differences between bvAD, typical AD and controls and significantly higher numbers in bvAD and typical AD compared to bvFTD. **Panel C** shows the ratio of total VENs (GABRQ-immunopositive and –negative) over all layer 5 neurons in order to correct for effects of neurodegenerative processes, showing no differences between bvAD, typical AD and controls in turn and significantly lower VENs in bvFTD than controls. **Panel D** shows the ratio of GABRQ-immunoreactive pyramidal neurons over all layer 5 neurons, showing no differences between bvAD, typical AD and controls and significantly higher ratios in bvAD and typical AD compared to bvFTD. Results were assessed using using emmeans post-hoc tests, adjusting for age, using a Bonferroni correction in R version 4.0.2.

## Discussion

In this study, we examined whether VENs and phylogenetically related neurons in the ACC show a selective loss in bvAD, potentially explaining the early and predominant behavioral changes that characterize this rare atypical variant of AD. Contrary to our hypothesis, we found no selective loss of VENs and GABRQ-ir neurons in bvAD compared to tAD and controls. These data suggest that, unlike in bvFTD, behavioral alterations in bvAD may not be related to a loss of VENs and phylogenetically related neurons in the ACC.

Our results are in line with a previous study showing no differences in the number of VENs of the ACC between bvAD and tAD [28] in patients with substantial coexisting α-synuclein pathology. In a sample of donors without comorbid neocortical α-synuclein nor TDP-43 pathology in both bvAD and tAD, our findings confirm the absence of a selective loss of VENs in the ACC in bvAD. Nevertheless, our results suggest a subtle, non-significant, trend towards intermediate levels of VENs in bvAD relative to tAD and bvFTD, while the previous study showed relatively higher levels of VENs in bvAD than tAD. In addition, we show no selective loss in bvAD in GABRQ-ir neurons, with significantly higher levels in bvAD compared to bvFTD. The discrepancy between the studies can be due to the differences in inclusion of donors with neocortical levels of α-synuclein pathology [28], where neocortical α-synuclein can also modulate the behavioral symptoms observed in bvAD. In our cohort, we aimed to include donors with limited copathologies, not reaching the ACC. More likely however, it could be due to the small sample size due to the rarity of this phenotype or due to the heterogeneous clinical and neuropathological presentation of bvAD[36].

Functional properties of VENs have been associated with intuitive decision making [37] in complex situations that are often characteristic of social situations [38]. VENs are located in the ACC and FI, which constitute key regions of the salience network [39] and regulate higher order social emotions like guilt, embarrassment and empathy as well as the subjective experience of pain and the need for autonomic activity [40], acting as a gateway between soma and psyche [41]. bvFTD shows specific vulnerability in these regions [42], leading to a wide range of social cognition deficits [43, 44]. Although social cognition deficits have been suggested in bvAD [45-48], reports are mainly based on case studies and possibly biased by cognitive deficits in bvAD. Therefore, future studies should incorporate larger groups and apply more objective measures of socioemotional functioning, such as biometric approaches [49] in order to capture the experiential component of social cognition. Based on the lack of loss of VENs and related neurons in the ACC and a lower involvement of salience network in bvAD than in bvFTD [22], social cognition patterns may show milder deficits in bvAD. Alternatively, VENs and related neurons may be more likely to show a selective loss in the frontoinsula in bvAD than in the ACC, as subtle frontoinsular hypometabolism was found previously [50] and no differences between bvAD and typical AD were observed in tau pathology within the ACC [26]. In addition, this study did not assess fork cells, which are also related to VENs and selectively targeted in bvFTD [2]. Future studies should explore these hypotheses.

Our finding that there is no reduction of VENs and related neurons in the ACC, combined with predominantly AD-typical patterns of neurodegeneration [12, 19, 20, 22] and the lack of consistent regional difference in tau pathology between bvAD and tAD [24, 26, 51] may suggest more diffuse neurobiological mechanisms to underlie the behavioral alterations in bvAD. Indeed, anterior default mode network [22], micro-level dysregulations, such as amygdalar overactivity[22] may contribute to its phenotype. Alternatively, developmental or premorbid personality structures that predispose individuals to exacerbation of vulnerable personality features with accumulating AD pathology. Therefore, future studies should aim to better characterize individuals with bvAD neurobiologically in terms of functional networks as well as clinically in terms of personality, coping styles, early life events, premorbid psychiatric conditions. Alternatively, bvAD may represent a heterogeneous phenotype, in which some patients may show neurobiological features similar to bvFTD and others may show a more “typical AD” neurobiological profile [52].

The current study should be evaluated in light of its limitations. First, the small sample size due to the rarity of the disease may have hampered statistical power. Second, copathologies are common in AD. Although we included donors with low levels of copathologies in bvAD and tAD, the potential contribution of these pathologies to their clinical manifestation is currently unknown. Second, the present study selectively sampled the ACC, while VENs and related neurons are also located in high numbers in the FI. Indeed, bvAD showed frontoinsular involvement based on patterns of atrophy [18, 53] and hypometabolism [22], while the ACC showed no differences in postmortem tau pathology in bvAD compared to tAD [26]. Alternatively, early preferential tau aggregation within VENs in the ACC could affect behavior, as has been observed in specific *MAPT* mutations leading to bvFTD [54]. In AD dementia, rare tau aggregations were identified in VENs, whose density was inversely correlated with Braak stage [55]. Future studies should investigate VENs in the FI and include early post-mortem cases in order to study early local aggregation of tau pathology within the ACC and FI in bvAD cases. Moreover, VENs and pyramidal neurons may not express GABRQ-immmunoreactivity, while actually being present, resulting in “phenotypic silencing”. In our data, we found no significant differences between groups in the number of GABRQ-negative VENs, GABRQ-negative pyramidal neurons or the percentage of GABRQ-negative VENs of all VENs (all *p*>0.05, Table S5 and Figure S1). In previous work, VENs were positively correlated with GABRQ-expressing neurons [10], suggesting that the loss of GABRQ-expressing neurons will be similar across groups. However, the possibility of unequal phenotypic silencing across groups cannot be ruled out.

In conclusion, our results demonstrate no selective loss of VENs and related neurons in the ACC in bvAD. Future research is needed to further elucidate the neurobiological mechanisms constituting the early and predominant behavioral dysregulation in bvAD.

## Data Availability

Data is available upon reasonable request.

