## Supplemental data for "The behavioral variant of Alzheimer’s disease does not show a selective loss of Von Economo and phylogenetically related neurons in the anterior cingulate cortex"

**Table S1.** Clinical characterization of behavioral features per Rascovsky criteria in patients with bvAD.

|  | bvAD #1 | bvAD #2 | bvAD #3 | bvAD #4 | bvAD #5 | bvAD #6 | bvAD #7 | bvAD #8 | bvAD #9 |
| --- | --- | --- | --- | --- | --- | --- | --- | --- | --- |
| Age | 70-75 | 70-75 | 60-65 | 75-80 | 55-60 | 60-65 | 55-60 | 75-80 | 80-85 |
| Sex | F | M | F | F | M | M | F | F | M |
| bvFTD symptoms |  |  |  |  |  |  |  |  |  |
| Disinhibition | Y | Y | Y | Y | Y | Y |  |  |  |
| Apathy |  | Y | Y |  |  |  | Y | Y | Y |
| Loss of empathy |  |  |  |  |  |  |  |  |  |
| Compulsiveness | Y | Y | Y |  |  |  |  | Y | Y |
| Hyperorality |  | Y | Y | Y | Y |  | Y |  | Y |
| Dysexecutive functioning |  | Y |  |  | Y | Y | Y | Y |  |
| Total | 2 | 5 | 4 | 2 | 3 | 2 | 3 | 3 | 3 |

**Table S2.** Demographic and pathological characteristics of participants by diagnostic group.

|  | bvAD | tAD | bvFTD-C9orf72 | bvFTD-sporadic | bvFTD-Progranulin | Controls | p-value |
| --- | --- | --- | --- | --- | --- | --- | --- |
| N | 9 | 6 | 9 | 4 | 5 | 13 |  |
| Age at death | 70.1 (9.0) | 79.7 (12.7) | 66.9 (8.5) | 65.3 (7.4) | 60.8 (10.6) | 66.8 (11.4) | 0.32 |
| Sex, no. of females, % | 5, 55.6 | 2, 33.3 | 4, 44.4 | 2, 50.0 | 4, 80.0 | 6, 50.0 | - |

†Significant differences in age were assessed by an ANOVA test and differences in sex were assessed with a χ^2^ test.

**Table S3.** Pathological characterization of copathologies in patients with bvAD and typical AD.

| # | Dx | Sex | | Age at death | | | | Disease duration (month) | | | Dx neuro-path. | | | ABC score | | | Brain weight (grams) | | | Cause of death | | | PMI (hrs) | | | CAA | | | CVD | | | | LB | | | TDP | | | ARTAG | |
| --- | --- | --- | --- | --- | --- | --- | --- | --- | --- | --- | --- | --- | --- | --- | --- | --- | --- | --- | --- | --- | --- | --- | --- | --- | --- | --- | --- | --- | --- | --- | --- | --- | --- | --- | --- | --- | --- | --- | --- | --- |
| 1 | bvAD | F | | 70-75 | | | | 13 | | | AD | | | A3 B3 C3 | | | 1230 | | | cardiac arrest | | | 15.0 | | | Absent | | | Unknown | | | | Absent | | | Absent | | | Present | |
| 2 | bvAD | M | | 70-75 | | | | 63 | | | AD | | | A3 B3 C3 | | | 1230 | | | cachexia | | | 3.42 | | | Present | | | Absent | | | | Amygdala | | | Absent | | | Present | |
| 3 | bvAD | F | | 60-65 | | | | 104 | | | AD | | | A3 B3 C3 | | | 860 | | | pneumonia | | | 7.17 | | | Present | | | Present | | | | Amygdala | | | Absent | | | Present | |
| 4 | bvAD | F | | 75-80 | | | | 120 | | | AD | | | A3B3C3 | | | - | | | dehydration | | | 4.55 | | | Present | | | Present | | | | Absent | | | Absent | | | Absent | |
| 5 | bvAD | M | | 55-60 | | | | 576 | | | AD | | | High-likelihood | | | 1192 | | | - | | | 5.50 | | | Present | | | Absent | | | | Absent | | | Absent | | | Absent | |
| 6 | bvAD | M | | 60-65 | | | | 96 | | | AD | | | A3B3C3 | | | 1260 | | | - | | | 8.50 | | | Present | | | Absent | | | | Absent | | | Absent | | | Absent | |
| 7 | bvAD | F | | 55-60 | | | | 72 | | | AD | | | A3B3C3 | | | 1030 | | | - | | | 17.5 | | | Present | | | Present | | | | Absent | | | Absent | | | Present | |
| 8 | bvAD | F | | 75-80 | | | | 48 | | | AD +some LB | | | A3B3C3 | | | - | | | dehydration | | | 7.30 | | | Present | | | Present | | | | Brainstem only | | | Absent | | | Present | |
| 9 | bvAD | M | | 80-85 | | | | 180 | | | AD | | | A3B3C3 | | | - | | | aspiration | | | 7.30 | | | Present | | | Present | | | | Amygdala | | | Limbic only | | | Absent | |
| *M* |  | | 55.6% F | | 70.1 (8.5) | | | | | 141.3 (160.0) | |  |  | | |  | | |  | | | 8.5 (4.5) | | | | | | 8/9 | | 5/8 | | 4/9 | | | 1/9 | | | 5/9 | | |
| 1 | tAD | | M | | | 85-90 | | | 48 | | | AD | | | A3B3C3 | | | - | | | dehydration | | | 8.45 | | | Present | | | | Present | | | Absent | | | Hippocampus only | | | Present |
| 2 | tAD | | M | | | 95-100 | | | 84 | | | AD | | | A3B3C3 | | | - | | | dehydration | | | 4.25 | | | Present | | | | Absent | | | Amygdala/Ob only | | | Absent | | | Present |
| 3 | tAD | | F | | | 65-70 | | | 34 | | | AD | | | A3 B3 C3 | | | 1065 | | | urosepsis | | | 6.17 | | | Present | | | | Present | | | Absent | | | Absent | | | Present |
| 4 | tAD | | F | | | 85-90 | | | 84 | | | AD + some LB | | | A3B3C3 | | | - | | | cachexia | | | 5.45 | | | Absent | | | | Present | | | Brainstem only | | | Limbic only | | | Absent |
| 5 | tAD | | M | | | 70-75 | | | >120 | | | AD | | | A3B3C3 | | | - | | | palliative sedation | | | 7.00 | | | Present | | | | Present | | | Absent | | | Absent | | | Present |
| 6 | tAD | | M | | | 60-65 | | | 120 | | | AD + some LB | | | A3B3C3 | | | - | | | dehydration | | | 7.55 | | | Present | | | | Present | | | Amygdala | | | Limbic only | | | Absent |
| *M* |  | | 33.3 % F | | | | 79.7 (11.6) | | | 81.7 (32.5) | |  |  | | |  | | |  | | | | | | 6.5 (1.4) | | | 5/6 | | 5/6 | | 3/6 | | | 3/6 | | | 4/6 | | |

**Table S4.** Numbers and ratios of VENs and GABRQ-immunopositive neurons in bvAD compared to tAD, bvFTD and controls.

|  | bvAD | tAD | bvFTD | CN | p-value, unadjusted | p-value, age adjusted | Post-hoc group differences unadjusted,  Bonferroni corrected | Post-hoc group differences with age adjustment,  Bonferroni corrected |
| --- | --- | --- | --- | --- | --- | --- | --- | --- |
| n | 9 | 6 | 18 | 13 |  |  |  |  |
| No. of VENs (mean, sd) | 26.00 (15.32) | 32.00 (18.05) | 10.94 (13.77) | 33.46 (20.27) | <0.001 | <0.001 | bvFTD<CN,  p<0.001 *** | bvFTD<CN,  p=0.003 **  bvFTD<tAD,  p=0.03 * |
| No. of GABRQ+ neurons (mean, sd) | 286.78 (101.16) | 380.83 (126.57) | 150.17 (91.47) | 370.31 (96.61) | <0.0001 | <0.0001 | bvFTD<CN,  p<0.0001 ****  bvFTD<bvAD,  p=0.01 *  bvFTD<tAD,  p<0.0001 **** | bvFTD<CN,  P<0.0001 ****  bvFTD<bvAD,  p=0.009 *  bvFTD<tAD,  p<0.001 *** |
| Ratio of VENs/all neurons (mean, sd) | 0.009 (0.004) | 0.012 (0.005) | 0.005 (0.005) | 0.010 (0.005) | 0.004 | 0.004 | bvFTD<CN,  p=0.01 *  bvFTD<tAD,  p=0.03 * | bvFTD<CN,  sp=0.02 * |
| Ratio of GABRQ+/all neurons (mean, sd) | 0.107 (0.022) | 0.144 (0.040) | 0.063 (0.031) | 0.116 (0.029) | <0.001 | <0.001 | bvFTD<CN,  p=0.0001 ***  bvFTD<bvAD,  p=0.005 **  bvFTD<tAD,  p<0.0001 **** | bvFTD<CN,  p=0.0001***  bvFTD<bvAD,  p=0.01 *  bvFTD<tAD,  p<0.001 ** |

*p<0.05, **p<0.01, ***p<0.001, ****p<0.0001. Abbreviations: *bvAD*=behavioral variant of Alzheimer’s disease; *bvFTD*=behavioral variant frontotemporal dementia; *CN*=cognitively normal; *tAD*=typical Alzheimer’s disease.

**Table S5.** Numbers of GABRQ-immunopositive VENs, GABRQ-immunonegative VENs, GABRQ-immunopositive pyramidal neurons and GABRQ-immunonegative pyramidal neurons per diagnostic group (Bonferroni corrected, age adjusted p-values)

|  | bvAD | tAD | bvFTD | CN | p-value, age adjusted | Post-hoc group differences age adjusted,  Bonferroni corrected |
| --- | --- | --- | --- | --- | --- | --- |
| n | 9 | 6 | 18 | 13 |  |  |
| No. of GABRQ-ir VENs (mean, sd) | 26.33 (15.90) | 31.00 (17.89) | 9.78 (13.53) | 30.92 (18.34) | 0.001 | bvFTD<CN, p=0.004 **  bvFTD<tAD, p=0.03 * |
| No. of GABRQ-negative VENs (mean, sd) | 1.00 (1.22) | 1.67 (1.97) | 1.17 (1.47) | 2.54 (3.36) | 0.16 | ns |
| No. of GABRQ-ir pyramidal (mean, sd) | 260.44 (87.13) | 349.83 (109.64) | 140.39 (82.58) | 339.38 (95.88) | <0.0001 | bvFTD<CN, p<0.0001 ****,  bvFTD<tAD, p<0.001 ***,  bvFTD<bvAD, p=0.01 * |
| No. of GABRQ-negative pyramidal (mean, sd) | 2442.22 (858.86) | 2514.33 (1543.91) | 2176.61 (767.65) | 2986.92 (772.35)◊ | 0.03 | ns |
| Percentage GABRQ-negative VENs of all VENs (mean, sd) | 5.75 (8.07)∆ | 4.32 (6.09) | 13.82 (14.00) | 5.88 (6.86) | 0.05 | ns |
| Percentage GABRQ-negative pyramidal of all pyramidal neurons (mean, sd) | 90.14 (1.94)∆ | 86.51 (3.74) | 94.08 (2.83) | 89.22 (2.99) | 0.0002 | bvFTD<CN, p<0.001 ***,  bvFTD<tAD, p<0.001 ***,  bvFTD<bvAD, p=0.02 * |

◊ n=12 for the negative pyramidal neurons in the CN group.
∆ n=15 for the percentage negative VENs and pyramidal neurons in the bvFTD group.

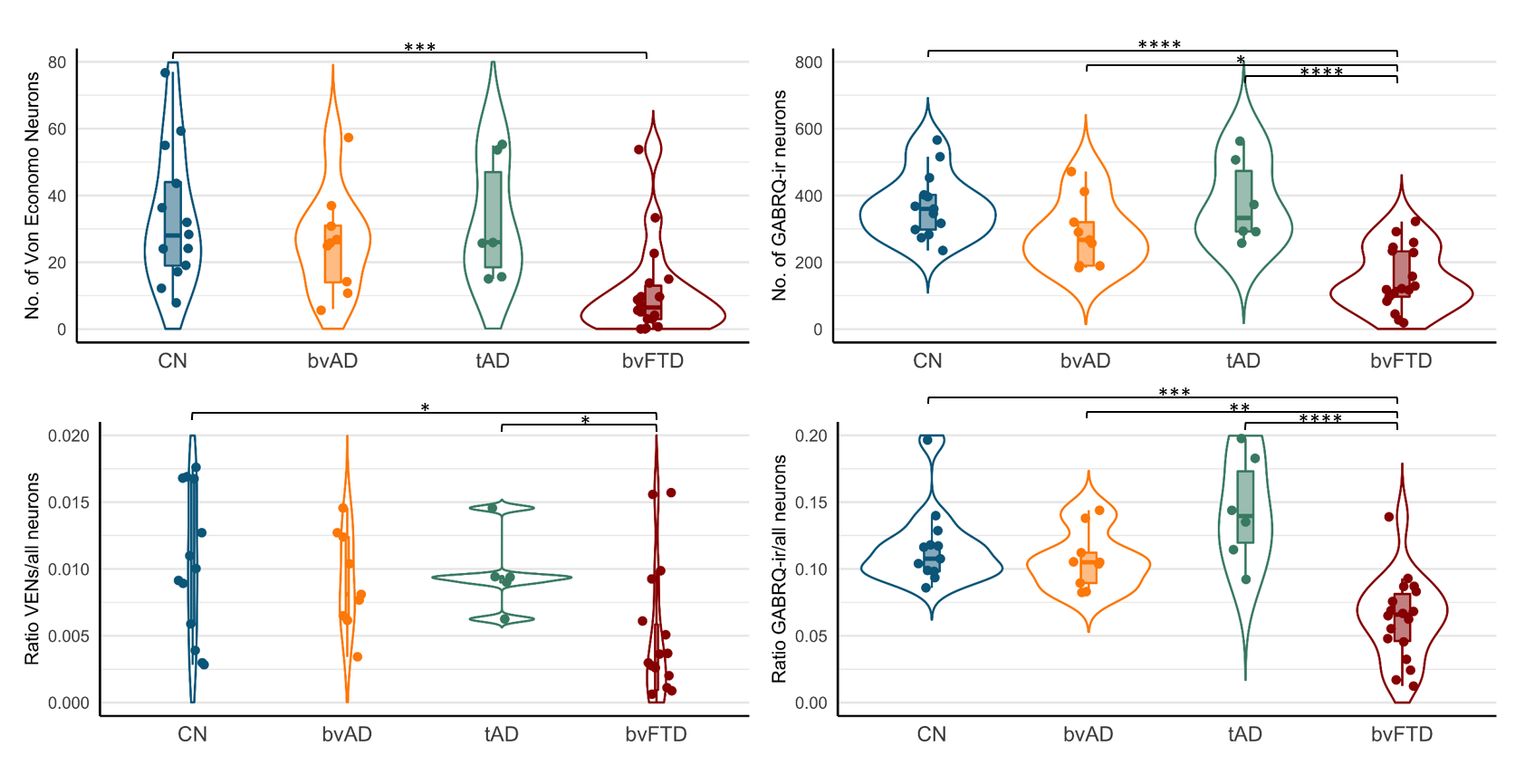
**Figure S1.** Numbers and ratios of VENs and GABRQ-immunopositive neurons in bvAD compared to tAD, bvFTD and controls (Bonferroni corrected, unadjusted p-values)

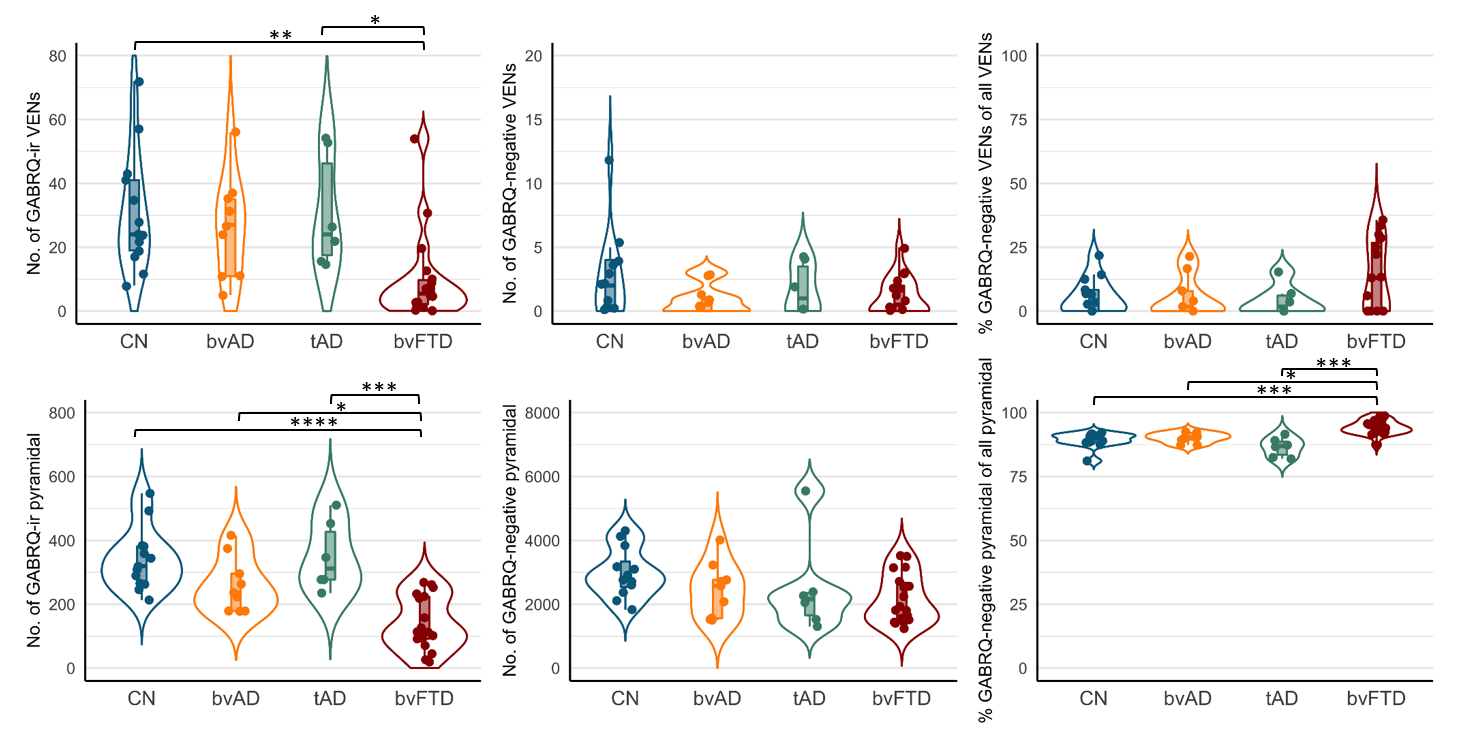
**Figure S2.** Numbers of GABRQ-immunopositive VENs, GABRQ-immunonegative VENs, GABRQ-immunopositive pyramidal neurons and GABRQ-immunonegative pyramidal neurons per diagnostic group (Bonferroni corrected, age adjusted p-values).

**Figure S3.** Plots showing the relationship between the number of bvFTD symptoms and number of VENs and GABRQ-pyramidal neurons in bvAD cases

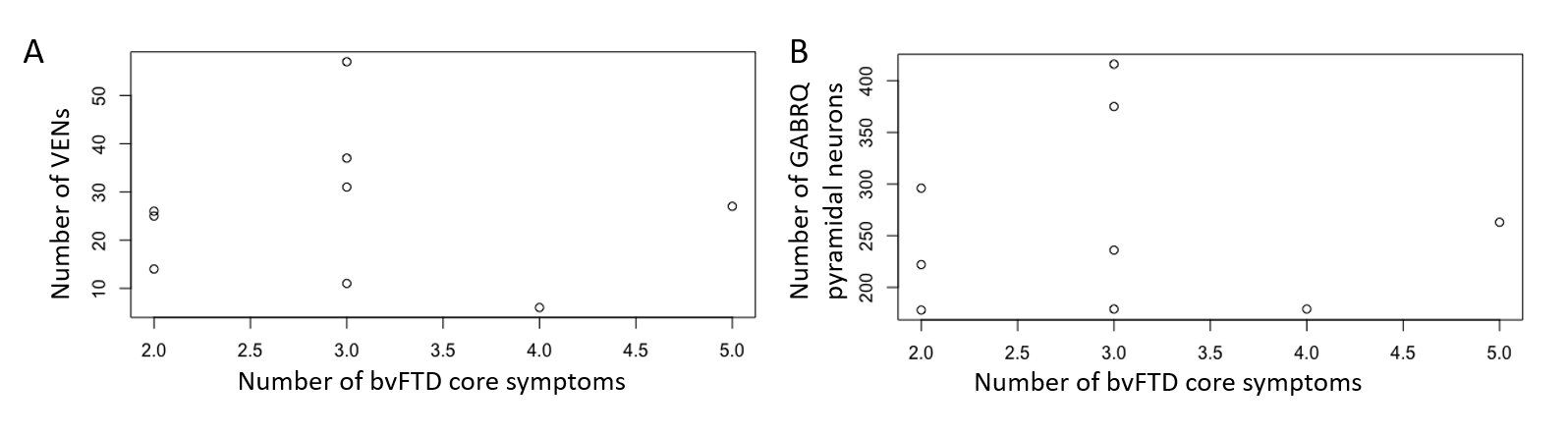
